# Causal Network Mapping of sEEG Identifies Compact Epileptogenic Targets Concordant with Seizure Freedom: A Multicenter Study of 97 Patients

**DOI:** 10.64898/2026.05.21.26353792

**Authors:** Alyssa Ailion, Alexander P. Rockhill, Hana Farzaneh, Rebecah Kaplun, Jeffrey Bolton, Melissa Tsuboyama, Alexander Rotenberg, Steven Lee, Abhay Dutta, Dekel Shapira, Darya Frank, Noam Peled, Paul Ferrari, Daniel J. Curry, Scellig Stone, Phillip L. Pearl, Howard L. Weiner

## Abstract

**Background and Purpose:** Drug-resistant epilepsy (DRE) affects approximately 15 million people worldwide, and surgery remains one of the only curative options. A key challenge in predicting outcomes is the lack of standardized, quantitative tools to help distinguish seizure driver regions from propagation regions during stereoelectroencephalography (sEEG) recordings. We validated a novel metric we call *criticality* which uses causal network mapping and machine learning to assign scores to sEEG contacts such that higher scores correspond to surgically treated tissue in patients with more favorable outcomes. Criticality is a per-contact score of how strongly each recording site drives the seizure network. Compared to the most similar previously published method, neural fragility, criticality also measures which nodes in the network have destabilizing (seizure-causing) network effects but also models time-delayed connections and frequency-specific interactions such as phase-amplitude and cross-frequency coupling.

**Methods:** We analyzed de-identified clinical data from 60 patients (aged ≥ 2 years) with focal or multifocal DRE who underwent sEEG monitoring and proceeded to surgery at four U.S. Level-4 epilepsy centers. The algorithm was trained on an independent cohort (N=37) and locked prior to validation. A random forest mapped each patient’s distribution of criticality values inside versus outside the treatment zone (TZ) to a probability of surgical success (P(success)), evaluated by leave-one-patient-out cross-validation. The primary outcome was the standardized effect size (Cohen’s d) of P(success) on the validation cohort between more favorable (Engel I–II) and less favorable (Engel III–IV) patient outcomes.

**Results:** The predicted P(success) was significantly higher in patients with more favorable outcomes in our held-out validation dataset (d = 1.12, 95% CI: 0.42–1.83, p = 0.001). Three potentially clinically actionable findings emerged: 1) High-criticality contacts formed spatially compact clusters (∼9 mm nearest-neighbor distance vs. 17 mm expected by chance), consistent with focal targets amenable to minimally invasive ablation. 2) Sensitivity was highest in small focal procedures (80% at ≤10 treated contacts) and decreased with resection size. 3) In patients with less favorable outcomes, high-criticality tissue remained outside the resection boundary, suggesting incomplete resection of the epileptogenic zone.

**Conclusions:** Our criticality metric applied causal network mapping to sEEG recordings, improving on the state-of-the-art benchmark for predicting treatment success probability in retrospective cases. Our criticality metric was most sensitive for focal, small-volume procedures and may be best suited for laser interstitial thermal therapy (LITT) and other minimally invasive approaches. When seizures persisted after surgery, residual high-criticality tissue outside the resection boundary offered both a mechanistic explanation for the less favorable outcome as well as potential targets for reoperation.

## 1. Introduction

Drug-resistant epilepsy (DRE), defined by the International League Against Epilepsy (ILAE) as the failure of two adequately trialed antiseizure medications [1], affects approximately one-third of the 65 million people living with epilepsy worldwide [2,3]. For these patients, surgery remains one of the only potentially curative treatments, yet success rates vary widely, from 60–70% in lesional temporal lobe cases to 30–40% in non-lesional or extratemporal cases [4,5,6,7,8]. Surgery that does not achieve seizure freedom carries potentially devastating consequences: continued seizures reduce quality of life and independence including progressive cognitive and behavioral decline [9], the risk of Sudden Unexpected Death in Epilepsy (SUDEP) is elevated [10], and there are sustained societal costs [11,12]. Surgeries that are successful at stopping seizures and that preserve cognitive abilities like language and memory (via pre-surgical function mapping [13]) allow patients to live healthier and more independent lives. Surgery is high stakes, and surgical planning to target an epileptogenic zone (EZ) is complex. Consider a common clinical scenario: a patient with non-lesional drug-resistant right frontal lobe epilepsy undergoes sEEG implantation. Their epileptologist reviews seven seizures each with slightly different timing of ictal onset in different recording channels. But which of those contacts are driving the seizures, and which are merely responding to propagated activity? This decision is typically made by expert visual analysis of the time series data. Expert analysis that does not quantitatively capture the directed network dynamics underlying seizure generation risks missing important information about seizure generation [14,15].

Contemporary research increasingly views epilepsy as a brain network disorder, where pathological network connectivity rather than a discrete lesion drives seizure generation and propagation [14,15]. Several computational approaches have been developed to interpret iEEG data. High-frequency oscillations (HFOs, 80–500 Hz), particularly ripples and fast ripples, represent the most extensively validated iEEG biomarkers for EZ localization; multicenter studies have demonstrated that HFO resection rates correlate with seizure outcomes [16,17,18]. Undirected functional connectivity metrics, including eigenvector centrality [19] and phase-locking values [20], have been applied to identify highly connected nodes, and graph-theoretic analyses have been used to map epileptogenic networks more broadly [21]. More recently, directed connectivity methods, including Granger causality [22], partial directed coherence [23], and information-theoretic approaches [24], have been used to infer causal relationships in seizure networks.

The only previous work we are aware of that validated the predictive value in terms of probability of a successful outcome for a patient using a multi-center cohort is neural fragility [25]. They showed that the fragility of nodes inside the Seizure Onset Zone (SOZ; the brain tissue rated by the patient’s epileptologist as causing seizures) compared to outside is far more predictive of the case outcome (favorable vs unfavorable) than frequency band-limited features such as HFOs. The HFO effect size, as measured by high-gamma-band activity (90–300 Hz), was much smaller than fragility’s (median Cohen’s d = 1.02 vs 1.51) [25]. Fragility is computed in short (125 ms) overlapping time windows, so it tracks how the network changes moment to moment rather than treating it as static. In each window, the method estimates how much each contact would need to be perturbed to push the network toward instability. Contacts that require only a small perturbation are ranked as the most “fragile,” yielding an interpretable, per-contact measure of which sites are most able to destabilize the network [25]. However, the estimation at each time point only depends on the time point sampled immediately previously, so fragility cannot capture delayed interactions; and because fragility does not separate interactions by frequency band, it may miss frequency-specific patterns of coupling that are relevant to seizure dynamics.

In this context, we report the clinical validation of a novel metric, criticality, that addresses these limitations. Criticality is based on delay-adjusted wavelet-based transfer entropy (dWTE) [24,26] computed on the sEEG recording. We used dWTE to measure not only whether two brain regions are related, but also whether activity in one region helps predict activity in another region. This makes it a directional measure of communication, helping identify which regions are putatively driving versus propagating seizure activity. Delay-adjusting allows for some delay for brain signals to travel from one region to another. Some interactions may occur with a time lag due to the distance neural signals must propagate; dWTE is designed to account for these propagation delays. Using wavelets allows the method to examine signal interactions over time and across different frequency ranges. The features from dWTE were optimized for decoding the probability of success using an XGBoost classifier [27], a machine-learning approach that combines many decision trees. This allows the classifier to learn complex patterns associated with seizure-related network activity from a large search space of features.

Our criticality metric can capture more complex patterns of communication between brain regions, such as relationships that are not simple or one-to-one (nonlinear synchrony), rhythms of activity that specifically drive communication (band-specific drivers), and differences in how quickly signals travel between pairs of regions (pair-specific conduction delays). For example, dWTE can distinguish fast communication between cortical regions from slower propagation through subcortical networks (fast cortico-cortical coupling versus slow subcortical propagation). In the fragility model, these features are largely forced into a more simplified framework: a linear model, broadband frequency information, and a single time scale for network interactions. As a result, criticality may capture a richer picture of how seizure-related activity moves through the brain. Our criticality metric can detect when activity in one frequency range influences or coordinates with activity in another frequency range, rather than assuming that each frequency band operates independently (i.e., detecting cross-frequency coupling). Because fragility relies on broadband frequency information, these frequency-specific relationships may be missed or compressed within the model.

This study aimed to validate that: 1) criticality identifies driver nodes in seizure networks that correspond to surgically treated tissue in patients with favorable outcomes, and 2) when seizures persist after surgery, untreated high-criticality tissue provides an explanation for the incomplete result.

## 2. Methods

### 2.1 Study Design and Setting

This retrospective, multicenter, observational, single-arm device-performance study utilized previously acquired de-identified clinical data. Data collection spanned January 2015 to December 2024. The four validation centers were the Hospital of the University of Pennsylvania (HUP), Johns Hopkins Hospital (JHH), University of Miami (UMF), and Texas Children’s Hospital (TCH). The training data were drawn separately from the Boston Children’s Hospital (BCH), the National Institutes of Health (NIH), and the University of Maryland Medical Center (UMMC), as described in Section 2.3. All validation cases used stereoelectroencephalography (sEEG) and/or electrocorticography (ECoG) recordings [28].

### 2.2 Participants and Eligibility

This study screened consecutive records from the retrospective epilepsy monitoring unit (EMU) registry. The inclusion criteria were as follows:

1. Patients aged ≥ 2 years
2. A diagnosis of focal or multifocal DRE
3. Surgical treatment with resection or ablation
4. Capture of at least three stereotyped seizures during sEEG/ECoG monitoring (for patients with more than three eligible seizures, all available stereotyped seizures were analyzed, and the per-seizure criticality scores were averaged to produce a single per-contact estimate of criticality)
5. Postoperative follow-up for at least 6 months with an established Engel outcome classification.
6. Availability of detailed operative notes or imaging explicitly listing resected or ablated electrode contacts.

Patients were excluded if they had a progressive neurodegenerative disease, or if they were missing operative notes that precluded the accurate spatial mapping of the treatment zone or missing pre- or postoperative structural imaging that was required for the analysis.

Of the 112 records screened across all sites, 5 were excluded for not meeting eligibility criteria and 10 for incomplete data, leaving 97 patients for the analysis.

### 2.3 Algorithm Training and Validation Split

To prevent data leakage, the XGBoost model was trained on a fully independent cohort of 37 patients before validation. This training set was derived from BCH (pediatric) and the NIH and UMMC subsets of the publicly available fragility multicenter dataset [25]. The validation cohort of 60 subjects did not overlap with the training cohort, and the two cohorts were drawn from entirely separate centers. The training cohort was used solely to derive the model weights and operating threshold; all performance claims were based exclusively on the independent validation cohort. The training cohort comprised 17 patients from BCH, 13 from NIH, and seven from UMMC; the validation cohort comprised 47 from HUP (drawn from the publicly available intracranial EEG dataset of Bernabei et al. [29]), nine from TCH, three from JHH, and one from UMF. In the training cohort, 19 patients were pediatric or young adult (≤ 21 years, referred to as pediatric hereafter) and 18 were adults, and 23 were male and 14 female. Cohort outcomes and recording types are summarized in Table 1; validation-cohort demographics are reported in Table 2 (Section 2.4).

**Table 1:** Study Cohorts, Outcomes, and Recording.

| Characteristic | Training (N=37) | Validation (N=60) |
| --- | --- | --- |
| **Contributing centers** | BCH (17), NIH (13), UMMC (7) | HUP (47), TCH (9), JHH (3), UMF (1) |
| **Outcome** |  |  |
| Favorable (Engel I–II) | 34 (92%) | 42 (70%) |
| Unfavorable (Engel III–IV) | 3 (8%) | 18 (30%) |
| **Engel Class** |  |  |
| Engel I | 31 (84%) | 34 (57%) |
| Engel II | 3 (8%) | 8 (13%) |
| Engel III | 2 (5%) | 12 (20%) |
| Engel IV | 1 (3%) | 6 (10%) |
| **Therapy** |  |  |
| Resection | 27 (73%) | 32 (53%) |
| Ablation | 8 (22%) | 28 (47%) |
| Not disclosed | 2 (5%) | 0 (0%) |
| **Recording** |  |  |
| sEEG | 17 (46%) | 39 (65%) |
| ECoG | 20 (54%) | 21 (35%) |
| **Treated contacts, median (range)** | 15 (4–89) | 10 (3–83) |

**Table 2:** Validation Cohort Demographics.

| Characteristic | Favorable<br>(N=42) | Unfavorable<br>(N=18) | Combined<br>(N=60) |
| --- | --- | --- | --- |
| **Age Group** |  |  |  |
| Adult (> 21 years) | 33 (79%) | 11 (61%) | 44 (73%) |
| Pediatric or young adult ($\leq 21$ years) | 9 (21%) | 4 (22%) | 13 (22%) |
| Not Disclosed | 0 (0%) | 3 (17%) | 3 (5%) |
| **Sex** |  |  |  |
| Male | 20 (48%) | 6 (33%) | 26 (43%) |
| Female | 22 (52%) | 9 (50%) | 31 (52%) |
| Not Disclosed | 0 (0%) | 3 (17%) | 3 (5%) |

### 2.4 Patient Demographics

A total of 42 patients achieved more favorable outcomes (Engel I/II), and 18 experienced less favorable outcomes (Engel III/IV). Outcome distribution did not differ significantly between pediatric (9/13 favorable, 69%) and adult (33/44 favorable, 75%) patients (two-tailed Fisher’s exact p = 0.73; age was undisclosed for the 3 JHH subjects). Sex data were unavailable for the 3 (5%) JHH subjects due to site-specific IRB de-identification protocols.

Lesion status, surgical target region, and implant laterality are summarized in Table 3. The cohort was split roughly evenly between lesional (37%) and non-lesional (40%) cases, with the remainder not disclosed by the contributing site. Targets were predominantly temporal, combining neocortical temporal (40%) and mesial temporal (25%) cases, with frontal (13%) and a small number of parietal, insular, and frontoparietal targets. Implant laterality is reported as the predominant hemisphere of implanted contacts derived from electrode coordinates; bilateral indicates substantial coverage in both hemispheres and does not denote bilateral seizure onset.

**Table 3:** Validation Cohort Clinical Characteristics.

| Characteristic | Favorable<br>(N=42) | Unfavorable<br>(N=18) | Combined<br>(N=60) |
| --- | --- | --- | --- |
| **Lesion status on MRI** |  |  |  |
| Lesional | 16 (38%) | 6 (33%) | 22 (37%) |
| Non-lesional | 18 (43%) | 6 (33%) | 24 (40%) |
| Not disclosed | 8 (19%) | 6 (33%) | 14 (23%) |
| **Surgical target region** |  |  |  |
| Temporal (neocortical) | 19 (45%) | 5 (28%) | 24 (40%) |
| Mesial temporal | 10 (24%) | 5 (28%) | 15 (25%) |
| Frontal | 4 (10%) | 4 (22%) | 8 (13%) |
| Parietal | 1 (2%) | 0 (0%) | 1 (2%) |
| Insular | 0 (0%) | 1 (6%) | 1 (2%) |
| Frontoparietal | 1 (2%) | 0 (0%) | 1 (2%) |
| Not disclosed | 7 (17%) | 3 (17%) | 10 (17%) |
| **Predominant implant hemi-sphere** |  |  |  |
| Left | 13 (31%) | 3 (17%) | 16 (27%) |
| Right | 9 (21%) | 3 (17%) | 12 (20%) |
| Bilateral | 20 (48%) | 12 (67%) | 32 (53%) |

### 2.5 Data Processing and Platform Architecture

Using ictal epochs (30 s pre- and post-seizure onset), the directed information flow between all contact pairs was computed using delay-adjusted wavelet-based transfer entropy (dWTE) [24,26]. This method quantifies whether activity at one contact helps predict activity at another, and in which direction. This is one method of causal network mapping. Directed connectivity values are fed into a locked machine-learning classifier (XGBoost [27]) that outputs a criticality score (0–1) for each contact: a score near 1 indicates a driver region that exports seizure activity to its neighbors, while a score near 0 indicates a propagation region or a brain area not involved in the seizure. For preprocessing: 1) Flat channels with peak-to-peak amplitude below 1 μV were removed, 2) the recording was down-sampled to 512 Hz, 3) power-line noise was removed with zero-phase infinite impulse response (IIR) notch filter, 4) For each annotated seizure, three clips were extracted relative to the electrographic onset: pre-ictal [−30, −5] s, peri-ictal [−5, +5] s, and post-ictal [+5, +30] s, 5) electrode contacts were bipolar referenced following the recommendation of Li et al. [30]. For each clip, and separately within each frequency band, we measured how much and in which direction activity at one contact predicted activity at another. The primary measure was a wavelet-based directed-connectivity method (multivariate phase-randomized wavelet transfer entropy, MPWTE), combined with two complementary measures of directional and cross-frequency interaction (the phase slope index, PSI, and phase-amplitude coupling, PAC). Signals were analyzed at frequencies centered on 4, 8, 13, 30, 55, 80, and 120 Hz. The resulting directed graphs were reduced to thirteen per-contact criticality scores: out-, in-, weighted-, and weighted-out strength; degree; clustering coefficient; betweenness, eigenvector, and closeness centrality; global- and local-efficiency drop on node removal; module-connector score; and out-strength-to-gamma. Each contact’s waveform shape was also summarized with catch22, a standard set of 22 time-series summary statistics [31]. For each contact, features were aggregated as the mean across clips within each window, band and metric triple, augmented by two cross-window differences 1) peri-ictal and pre-ictal, 2) post-ictal and pre-ictal, yielding 438 features. These graph and waveform metrics were assembled as an exploratory, data-driven feature set, and we did not pre-specify which of them would be informative. Rather than testing each feature for significance, all features were passed to the machine-learning classifier. Overfitting and false positives were controlled by locking the model and its threshold on the independent training cohort and evaluating them only once on the held-out validation cohort. Cohen’s d was obtained by patient-level bootstrapping.

### 2.6 Clinical Ground Truth and Outcome Anchoring

The electrode contacts that were treated were determined from postoperative T1 imaging by segmenting the resection/ablation area aided by the operative notes. Previous work has used Seizure Onset Zone (SOZ) to denote areas designated by the epileptologist as the brain tissue starting seizures [25]. We used the segmentation of this Treatment Zone (TZ) as the ground truth instead since it is objective. Healthy tissue is often removed during larger open surgical resection because of the surgical technique (i.e., temporal lobectomy), so the TZ is often a superset of the minimal brain area required to cure epilepsy. The algorithm could label some healthy tissue that was removed as false positives because of our choice of TZ as ground truth. We evaluated the algorithm’s performance based on the size or extent of surgical intervention to assess the effect of using TZ as ground truth.

The multicenter fragility dataset does not provide a segmented TZ, so for patients drawn from it the clinician-annotated SOZ was used as a fallback for the TZ. This affected 4 of the 60 validation patients and 20 of the 37 training patients; all other patients had a TZ defined from post-operative resection or ablation records. The implications of this substitution are addressed in the Limitations.

Surgical outcomes were assessed using the 6-month Engel classification [32]. More Favorable Outcomes were defined as Engel Class I (complete freedom from disabling seizures) and Engel Class II (rare disabling seizures). The inclusion of Engel II is clinically justified, as transitioning to rare seizures represents a substantial reduction in seizures, an improvement in overall quality of life, and a reduction in psychosocial morbidity compared to a less favorable outcome (Engel III–IV) [33]. The Engel score assignment was performed by epileptologists at each clinical site as part of routine clinical care. The scorer was blinded to the criticality outputs. All surgical decisions were made independently of and prior to this analysis.

### 2.7 Statistical Analysis

The primary outcome was the standardized effect size (Cohen’s d) of the output of the algorithm (the estimation of the probability of treatment success between more favorable and less favorable patient outcomes) using the criticality values as input. The between-group Cohen’s d was reported with a bias-corrected and accelerated (BCa) bootstrap 95% confidence interval (5,000 resamples) [34], accompanied by a two-sided Mann–Whitney U test p-value and the area under the ROC curve (AUC).

P(success) was produced by a random-forest classifier distinct from the criticality model. For each patient, we summarized how high the criticality scores were inside the treatment zone (TZ) and, separately, outside it. We did this by taking ten evenly spaced summary values (the 10th through 100th percentiles) of the scores in each region, giving 20 features per patient. A random forest (500 trees, balanced class weights) mapped these features to a patient-level probability of a favorable (Engel I–II) outcome, and per-patient P(success) was estimated by leave-one-patient-out cross-validation, each patient scored by a forest fit on all others. Note that the criticality model was trained and locked on the independent cohort but this probability of success estimation step uses cross-validation on the validation cohort following the approach of [25].

Because the sample size (N = 60) and the outcome split (42 favorable, 18 unfavorable) were fixed by the retrospective cohort, we report the power this sample affords rather than a prospective sample-size calculation. At α = 0.05, the sample provides approximately 10% power to detect a small effect (Cohen’s d = 0.2), 42% power for a medium effect (d = 0.5), and 80% power for a large effect (d = 0.8) [35].

Contact-level sensitivity, specificity, PPV, and NPV were evaluated in favorable-outcome patients only, where the treatment zone serves as a validated approximation of the epileptogenic network. A threshold for high criticality was selected by maximizing Youden’s J index (sensitivity + specificity − 1) [36] on the training cohort. Criticality scores are normalized by this threshold (divided by 0.15) so that values above 1 denote high-criticality contacts. This was to make visualization more intuitive and for contact-level analyses that required binarization of the criticality values.

Spatial compactness of high-criticality contacts (threshold ≥ 0.15) was assessed from per-contact Euclidean coordinates derived from co-registered imaging. For each patient we computed the mean nearest-neighbor distance among high-criticality contacts and the clustered fraction (proportion with at least one high-criticality neighbor within 10 mm), and compared these to a per-patient permutation baseline (2,000 random relabelings); observed-versus-expected differences across patients were tested with the Wilcoxon signed-rank test. This tests whether the contacts the algorithm flags as most critical sit close together in a small region of the brain, as would be expected for a focal target amenable to resection or ablation, rather than multiple distributed foci.

To test whether continuous criticality scores carry rank-order information beyond the binary above/below-threshold classification, the criticality values were related to TZ membership within the high-criticality subset (946 contacts in 42 patients): contacts were divided into score deciles, the proportion inside the TZ (naïve pooled PPV) was computed per decile with Wilson 95% confidence intervals, and the score–membership association was quantified by Spearman’s rank and point-biserial correlation.

## 3. Results

### 3.1 Probability of Success Analysis

On the validation cohort, P(success) separated the two outcome groups: mean P(success) was 0.78 in favorable (Engel I/II) patients versus 0.54 in unfavorable (Engel III/IV) patients (Cohen’s d = 1.12, 95% CI: 0.42–1.83; Mann–Whitney p = 0.001; AUC = 0.77). At the Youden-optimal operating threshold (0.86), P(success) classified surgical outcome with 50% sensitivity and 94% specificity (Table 4).

| Favorable mean<br>$P(\text{success})$<br>($N=42$) | Unfavorable<br>mean $P(\text{success})$<br>($N=18$) | Cohen's $d$ | p-value | Sens-<br>itivity | Spe-<br>ci-<br>ficity | AUC |
| --- | --- | --- | --- | --- | --- | --- |
| 0.78 | 0.54 | 1.12 (95%<br>CI 0.42–<br>1.83) | 0.001 | 50% | 94% | 0.77 |

In an exploratory subgroup analysis, P(success) separated more favorable from less favorable outcomes in the same direction for both lesional (n=22; Cohen’s d = 0.67, AUC = 0.72) and non-lesional (n=24; Cohen’s d = 0.94, AUC = 0.69) cases, indicating comparable discrimination across the two groups. These subgroups were small (six less-favorable patients each), so neither reached significance alone (Mann–Whitney p = 0.12 and p = 0.20), and lesion status was undisclosed for the remaining 14 patients.

### 3.2 Illustrative Case

To illustrate how our criticality metric translates to potential clinical decision-making, we present a representative case from the validation cohort in which the criticality map is used to weigh alternative treatment plans against their model-estimated probability of success, P(success). The bipolar contact criticality map is displayed alongside the electrode contacts with three candidate surgical treatment plans (Figure 1a–c). For each plan, the locked model returns a P(success) for the proposed treatment volume, allowing the surgical team to reason about the added value of treating each additional brain area against the risk of functional or cognitive deficits.

**Figure 1a.**
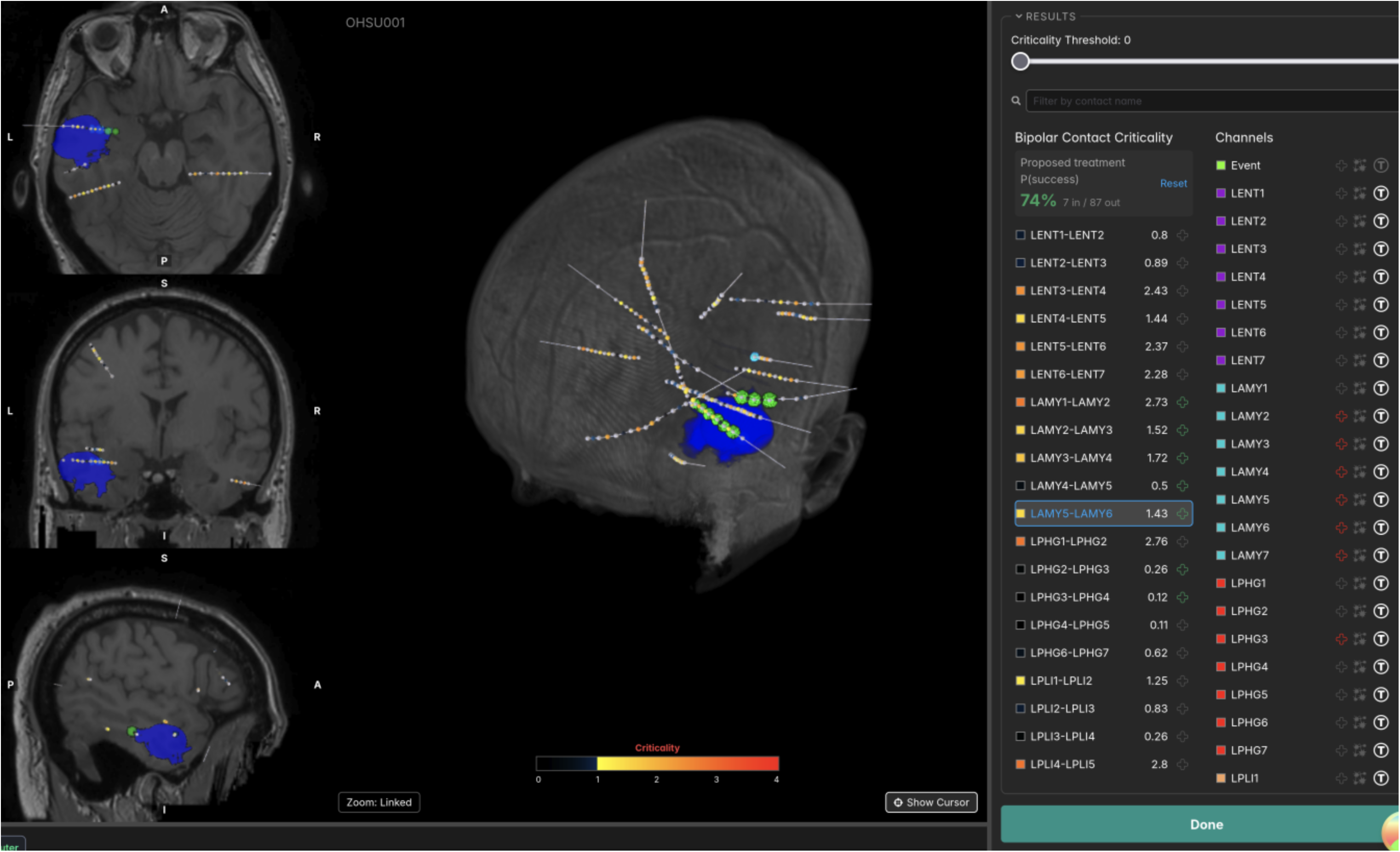
Criticality-guided treatment planning for an illustrative case: the treatment actually performed. This treatment resulted in an Engel IIA outcome; the model estimates P(success) = 74% for this treated volume. The left column shows co-registered MRI slices, the center shows the 3D implant with the criticality colormap, and the right lists per-contact criticality scores and the contacts included in plan. Criticality values are shown normalized by the threshold determined by Youden’s J analysis.

**Figure 1b.**
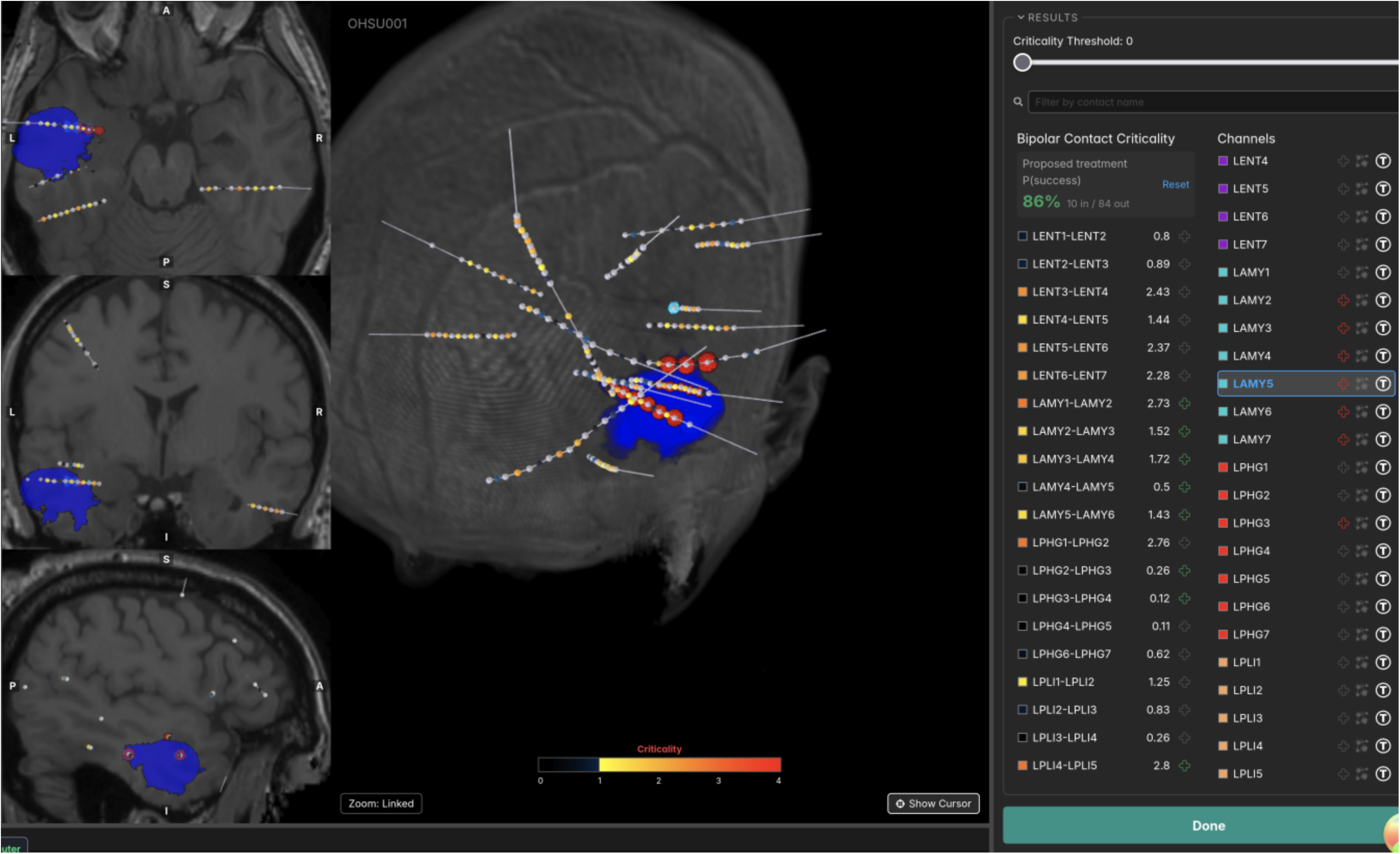
Superior expansion of the resection. Expanding the resection superiorly to capture additional high-criticality tissue raises the model-estimated P(success) to 86%. Columns are as in Figure 1a.

**Figure 1c.**
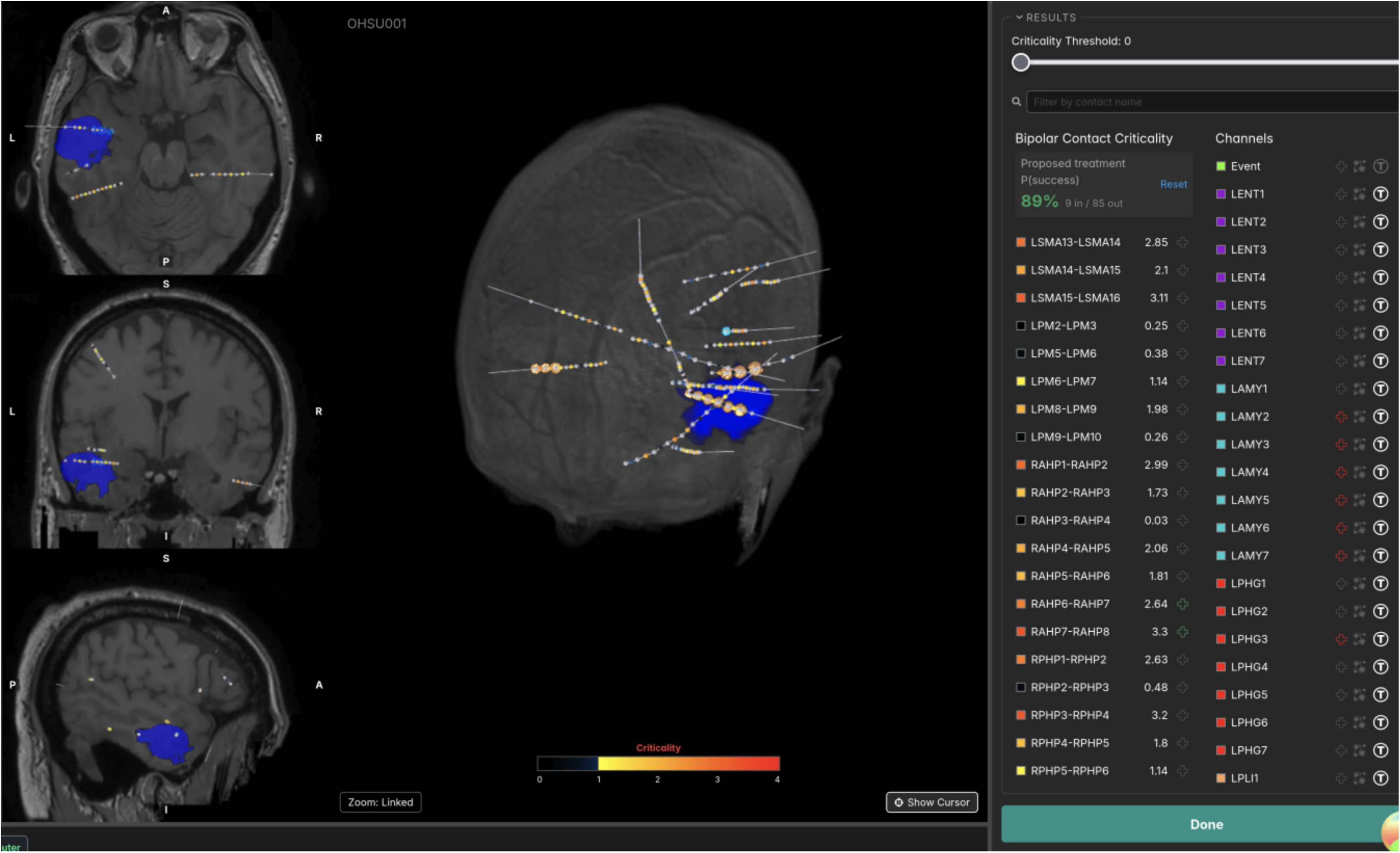
Additional contralateral-hippocampus target. Targeting the contralateral hippocampus in addition raises P(success) to 89%, but carries a substantial risk of cognitive deficit. Columns are as in Figure 1a.

### 3.3 Treatment-Zone Criticality Enrichment

The difference in P(success) between patients is driven by what happens at the level of individual contacts: criticality scores were higher on contacts inside the treatment zone than on contacts outside it. Across all 5,611 validation contacts, the mean criticality score was 0.28 inside the TZ versus 0.10 outside it (Cohen’s d = 0.97, Mann–Whitney p = 2.3 × 10⁻⁸⁹). Using the locked high-criticality threshold to flag TZ membership yielded 49% sensitivity and 83% specificity (Table 5), consistent with the contact-level performance reported in Section 3.4.

**Table 5:** Treatment-Zone Criticality Enrichment (validation cohort, 5,611 contacts)

| Metric | Value |
| --- | --- |
| Contacts inside TZ (n) | 912 |
| Contacts outside TZ (n) | 4,699 |
| Mean criticality inside TZ | 0.28 |
| Mean criticality outside TZ | 0.10 |
| Cohen's $d$ | 0.97 |
| p-value (Mann–Whitney) | $2.3 \times 10^{-89}$ |
| Sensitivity (high-criticality → in TZ) | 49% |
| Specificity (high-criticality → in TZ) | 83% |

### 3.4 Treatment-Zone-Size Analysis

Patients with larger resections tended to have lower sensitivity: per-patient sensitivity fell as the number of treated contacts increased (Spearman’s rho = −0.62, p < 0.0001). In the smallest stratum (≤ 10 treated contacts, n=21, representing 50% of the favorable cohort), the mean sensitivity was 80%, whereas the sensitivity fell below 40% in patients with more than 30 treated contacts. Specificity moved in the opposite direction, tending to be higher in patients with larger resections (rho = 0.43). For context, the training cohort (BCH/NIH/UMMC, N=37) had a mean of 20.76 treated contacts per patient (median 15, range 4–89), confirming that the model was trained across a broad spectrum of contact burdens and was not optimized exclusively for small focal cases.

Bucketed analysis by treated-contact burden (groups of 10 contacts) confirmed this gradient (Figure 2), providing a practical interpretation rule: the algorithm is the most informative for focal procedures and should be read with increasing caution as the resection size increases.

**Figure 2.**
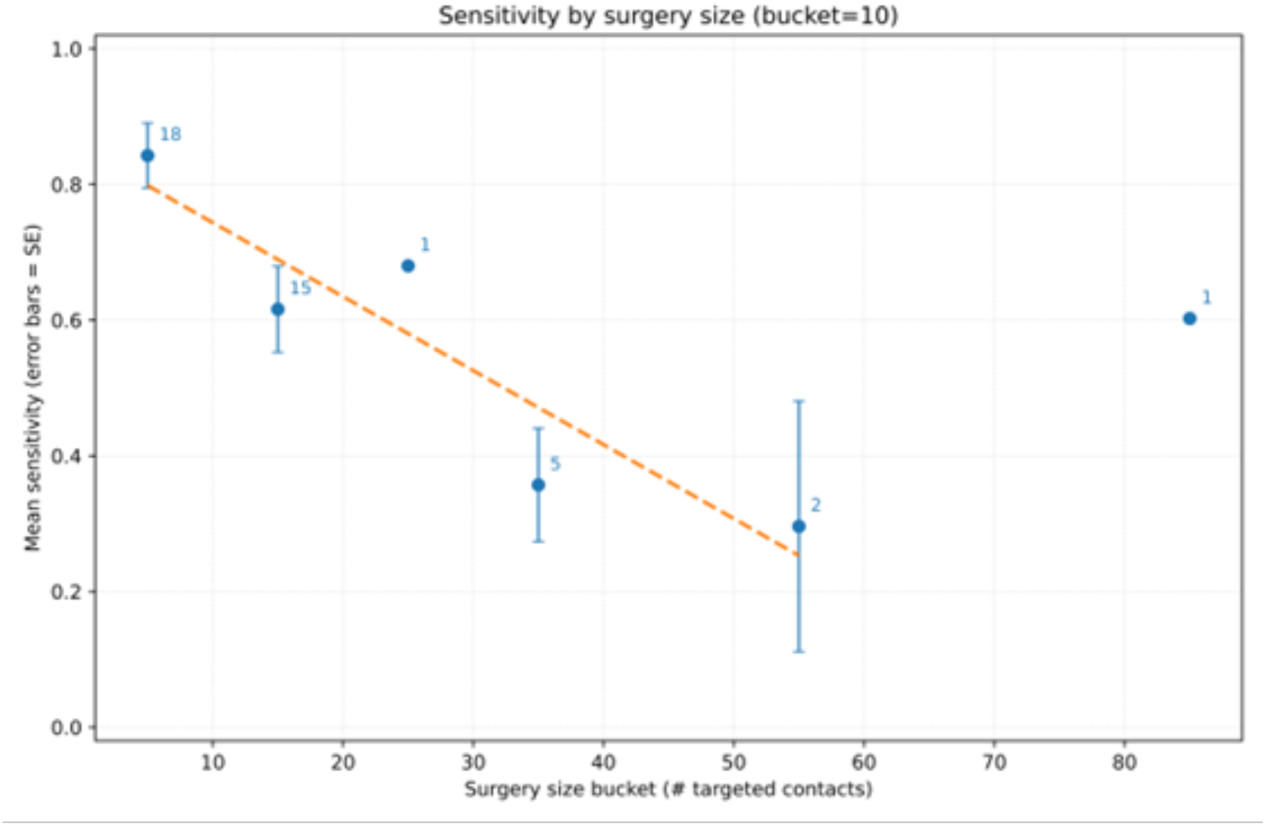
Mean per-patient sensitivity by surgery size bucket (10-contact bins, threshold 0.15, Engel I/II cohort, n=42). Error bars indicate standard error. The integers above each point indicate the number of patients per bucket. The trend line excludes the single-patient bucket at ≥ 80 treated contacts.

### 3.5 Spatial Compactness of Identified Driver Networks

A central question for surgical planning is whether the algorithm’s high-criticality contacts form targetable focal subnetworks or are diffusely distributed across the implanted field. Of the 42 patients, eight had missing coordinate files and were therefore excluded; these exclusions were driven by site-level data availability and were unrelated to the algorithm’s performance.

Among the 34 analyzable patients, the contacts flagged as most critical tended to lie in contiguous or closely adjacent cortex rather than distributed uniformly; they were substantially more spatially compact than expected by chance. The mean nearest-neighbor distance was 8.96 mm observed versus 17.21 mm expected (Wilcoxon p = 1.16 × 10⁻¹⁰), and the clustered fraction within 10 mm was 0.77 observed versus 0.39 expected (p = 3.20 × 10⁻⁹).

Within the high-criticality subset, criticality scores were significantly associated with TZ membership (Spearman’s rho = 0.28, p = 6.80 × 10⁻¹⁹; point-biserial r = 0.29, p = 1.28 × 10⁻¹⁹), indicating that higher criticality scores identify tissue with a progressively greater likelihood of being epileptogenic.

### 3.6 Mechanistic Analysis of Less Favorable Outcomes

To understand why some surgeries do not achieve seizure freedom despite iEEG-guided planning, we compared the distribution of high-criticality contacts relative to the treatment zone across the outcome groups (n=42 favorable, n=18 unfavorable). In patients with less favorable outcomes, the algorithm flagged high-criticality contacts outside the treated region. Tissue identified this way is not always removable; it may overlap eloquent cortex, carry unacceptable vascular risk, or be left in place after multidisciplinary and family discussion.

To quantify this, for each patient we compared the criticality scores on treated contacts with those on untreated contacts and measured how clearly the two groups could be separated (n=59 analyzable; one patient was excluded for < 2 contacts outside the treatment zone). Treating the Engel outcome as an ordinal score (I–IV), within-patient separation was inversely associated with Engel class, both when separation was measured as the difference in mean criticality between treated and untreated contacts (delta-mean; Spearman’s rho = −0.31, p = 0.016) and as the per-patient AUC distinguishing treated from untreated contacts (rho = −0.23, p = 0.083). Greater separation therefore tracked with better outcomes, a relationship driven mainly by the contrast between favorable (Engel I–II) and less favorable (Engel III–IV) classes rather than a strict gradient across all four.

The potential clinical implication is that before surgery, the epileptologist or surgeon could examine the proportion of high-critical contacts that fall within versus outside the proposed resection boundary. A large amount of untreated high-criticality tissue may serve as a preoperative warning that the planned intervention is insufficient to capture the entire driver network.

### 3.7 Analysis of Confounds

The validation cohort had a site imbalance, with the HUP accounting for 78% of the subjects. JHH contributed only three patients, all with unfavorable outcomes, precluding site-specific effect size estimation; this reflects consecutive registry sampling (eligible patients enrolled in a sequence at each site without outcome-driven exclusions) rather than systematic selection bias. An excluding-HUP sensitivity analysis (n=13) yielded d = 0.68 (CI: 0.12–1.24), suggesting, but not definitively establishing, that the effect is not driven by the HUP alone, given the limited power.

## 4. Discussion

This multicenter validation demonstrates that causal network mapping of sEEG provides three pieces of information that can potentially aid decision-making in an epilepsy surgery conference:

1. Statistical evidence of how likely a surgery is to succeed. The algorithm reliably indicates how likely a patient is to have a favorable or unfavorable outcome.
2. Evidence that the most favorable outcomes followed surgery on a more spatially restricted seizure onset zone. Criticality values produce a spatial map showing where the seizure drivers are, and that they cluster in compact neighborhoods that average 9 mm in size, which is well within the targeting precision of laser interstitial thermal therapy (LITT) [37,38,39]. Higher criticality scores mean a higher likelihood of epileptogenic tissue, while regions the algorithm scores as non-critical are very unlikely to harbor seizure drivers, providing reassurance when deciding where to stop resecting.
3. A warning that high-criticality contacts left outside the planned resection carry a risk of a less optimal outcome. When critical tissue falls outside the proposed resection boundary, that is a warning signal in our data; this pattern characterized the less favorable outcomes.

Taken together, these three outputs map onto concrete decisions in the surgical conference. Before surgery, the team can weigh the estimated likelihood of a favorable outcome, see whether the seizure drivers form a single focal target amenable to a minimally invasive approach such as LITT or a larger resection, and check whether any high-criticality contacts fall outside the planned treatment. When high-criticality tissue lies outside the plan, this can prompt a reappraisal of the resection or ablation strategy, or, where that tissue cannot be safely treated, a frank preoperative discussion of the reduced chance of seizure freedom. In patients who continue to have seizures after surgery, the same map highlights residual driver tissue as a candidate target for reoperation.

## 5. Limitations

The primary limitation of this study was its retrospective design. Although this ensured no risk to subjects and provided an objective, outcome-anchored validation framework, prospective integration into multidisciplinary epilepsy surgery conferences is an important future direction to measure the platform’s direct impact on real-time decision-making.

Data were collected from 2015 to 2024, a span over which recording systems, neuroimaging, and surgical techniques advanced considerably. These advances are potential temporal confounders: patients treated later in the period may have benefited from improved imaging and less invasive approaches, which could independently influence outcomes and therefore the apparent performance of the algorithm. The pre-locked threshold of 0.15 may not be optimal for all clinical settings; thus, prospective studies should evaluate adaptive thresholding.

Our cohort combined depth (sEEG) and subdural (ECoG) recordings, which sample the brain differently. These differences in spatial sampling could influence the connectivity estimates and the spatial-compactness metrics, and criticality has not yet been separately validated within each recording type; modality-specific prospective evaluation is warranted. Implant density of sEEG and ECoG varied between patients and hospitals, so using as large of a dataset as possible, especially with data from many different hospitals, is of utmost importance.

Patients from the multicenter fragility dataset were labeled by the clinician-annotated SOZ rather than a segmented TZ. SOZ and TZ are typically disjoint sets; some brain areas are taken during the surgery as margin or to access the epileptogenic tissue that are not labeled SOZ and some SOZ areas are to risky to treat because doing so may cause worse deficits that would be caused by the seizures, but, in general, TZ and SOZ largely overlap (median 60% overlap for the HUP dataset with both anotations). Because about half of the training cohort used SOZ, the results are more difficult to interpret. However, in other work, we compared SOZ to TZ using the fragility algorithm on the public fragility dataset as well as the dataset presented in this paper; the TZ results were comparable to SOZ with TZ having an edge in performance (AUC 0.59 for TZ versus 0.52 for SOZ) [40].

The 3D clustering analysis was limited to 34 of the 42 favorable patients because of missing coordinate data at certain sites. These exclusions were driven by site-level data availability and were unrelated to the algorithm performance. This outcome-pattern analysis was retrospective and explanatory; outcome labels were unavailable at the time of inference and should not be interpreted as predictive features. Prospective validation of distribution-distance metrics as indicators of preoperative coverage adequacy is warranted.

A more fundamental limitation is that criticality is computed relative to the implanted contacts and therefore ranks only the tissue that was sampled. A high criticality score identifies the most driver-like contacts among those recorded, but it cannot by itself confirm that the implant captured the true epileptogenic zone. If the seizure drivers were never sampled, the highest-criticality contacts may themselves be followers, and the resulting map could be misleading. The method does not yet provide an absolute criticality threshold, a minimum number of high-criticality contacts, or a network-topology criterion that certifies coverage adequacy. Criticality is therefore best interpreted after the clinical team has judged, from the seizure onset pattern and other data, that the implant plausibly sampled the epileptogenic zone; developing quantitative indicators of coverage adequacy is an important direction for future work.

Some race, ethnicity, and sex data were unavailable because of institutional deidentification protocols. There is no known biological mechanism by which demographic factors alter sEEG-based network connectivity. Among the 57 patients with available sex data, no significant difference in the treatment zone enrichment ratio was observed between the male (n=26) and female (n=31) sub-groups, although the comparison was underpowered. The validation cohort was drawn exclusively from U.S. Level-4 centers; applicability to lower-volume centers, non-U.S. settings, subdural grid recordings, or non-resective interventions such as neuromodulation, requires prospective evaluation.

## 6. Conclusion

Criticality identifies compact epileptogenic driver networks within the broader iEEG field and provides surgeons with a quantitative, reproducible map that can potentially inform precise surgical targeting. In this retrospective multicenter validation, the platform demonstrated a significant group-level separation between the outcome groups (d = 1.12, 95% CI: 0.42–1.83, p = 0.001).

## Ethics Statement

This study utilized existing de-identified clinical data and was exempt from Institutional Review Board (IRB) review under 45 CFR 46.104(d)(4). No patient consent was required as no identifiable private information was obtained. All data were handled in accordance with the institutional data-use agreements between FIND Neuro and the participating epilepsy centers.

## Data Availability

The clinical data and analysis will be made available upon request subject to a data use agreement (DUA) with each hospital in the study.

